# Impact of COVID-19 pandemic on use of Pediatric Emergency Health Services in a Tertiary Care Pediatric Hospital in North India

**DOI:** 10.1101/2021.01.09.21249489

**Authors:** Ravitanaya Sodani, Shalu Gupta, Virendra Kumar

## Abstract

**Objective:** To compare Pediatric Emergency attendance pre-COVID 19 to that during COVID 19 pandemic and to study changes in patient profiles attending Pediatric Emergency Department during COVID 19 pandemic.

**Methods:** We conducted a retrospective cross-sectional observational study and collected data from Medical Record Section during the COVID-19 pandemic from January to June 2020 and compared it with data from 2019 in similar months. Data collected was analyzed to find out the impact of COVID – 19 on use of pediatric emergency health services with respect to patient attendance, age and clinical profile before and during COVID-19 in a tertiary care hospital in New Delhi.

**Results:** We observed a 43% decline in PED visits which increased to 75% during the period of lock-down (p value = 0.005). There was a significant decrease in children of age group 1-5 years attending PED. Mortality rate during lockdown had gone up by nearly 3times than the average monthly mortality.

**Conclusions:** While children might not have been directly affected by the COVID-19 pandemic, but the fear of COVID 19 and measures taken to control the pandemic has affected the health seeking behavior of patients to an extent that indirectly caused more damage than anticipated.

## Introduction

On the 30^th^of January 2020, the World Health Organization declared the novel coronavirus outbreak as a public health emergency of international concern. Since the detection of first case of Coronavirus disease (COVID-19) in India on January 30, 2020, there was a rapid increase in number of COVID-19 positive cases. Almost instantaneously, the severe acute respiratory syndrome coronavirus 2 (SARSCoV-2) spread to all parts of the country penetrating the largest of towns, the smallest of villages and everything in between. As a response to the outbreak, various measures were implemented to prevent and contain the spread of infection. A nationwide lockdown was declared on the 24^th^of March 2020, which lasted till 30^th^ May followed by a staggered unlock process. However, the implications of the pandemic and its subsequent measures were unanticipated. COVID-19 has adversely affected the routine preventive and curative services especially for people with chronic conditions and disabilities.

In order to find the impact of the COVID-19 pandemic on use of health services – especially emergency services – we compared patient’s attendances in OPD & Pediatric Emergency Department (PED) and the changes in clinic epidemiological profiles (before and during COVID-19) in a tertiary care hospital in New Delhi.

### Study Design and Participants

We conducted a retrospective observational study in a tertiary care hospital (395 beds) in New Delhi, India with an average yearly OPD attendance of 2.8 lakh patients and 30,000 admissions (pre-COVID 19). The hospital has 24 hr. working Pediatric Emergency with nearly 1500 average Emergency admissions every month (Pre-COVID 19). Patient data of OPD attendance and of PED visits from January to June 2020 was retrieved from Hospital Medical Record Section. We compared Pediatric OPD attendance and Emergency attendance pre-COVID 19 and during COVID 19 pandemic. The study studies the trends for the periods from January to June 2020 in relation to the same months in 2019. We also compare patient profiles and outcome in terms of death or discharge/transfer attending Pediatric Emergency Department before and during COVID 19 pandemic. All patients attending the Pediatric OPD and PED were included in the study while patients with incomplete records were excluded from the analysis. The study has been reviewed and approved by Institutional Ethics Committee. Descriptive data was compared on statistical parameters such as means and standard deviations. Continuous data was analyzed using-test’s (p value significant reporting at <5%). Percentage changes were calculated to highlight differences in various patient parameters under consideration.

## Results

### Impact of COVID 19 on Hospital Attendance

The total hospital attendance, during the study period (January to June 2020), was 65,421individuals: of which 15,432 attended PED. Out of those who attended PED, 5,143 were admitted and 40 expired within 24 hours of admission.

On comparing data between 2020 and 2019, we found that routine OPD attendance reduced by 56%. Pediatric Emergency Service usage reduced from a monthly average of 4,500 in year 2019 to 2,572 in 2020: a 43% decline. Average daily attendance in PED reduced from 166 in 2019 to 85 (−48%) in 2020.We also observed that the average hospital admissions drastically reduced during COVID-19 by nearly 30%.(Table 1) This decline in PED attendance was more pronounced during the period of lockdown when this indicator decreased to nearly 75%. This decrease in PED attendance was statistically significant for the months – April, May and June (p-value = 0.005). On comparing the age profiles in the two study periods, we found that there was a significant decrease in children aged 1-5 years attending PED. Figure 1 shows the trend in PED admissions and death in 2020 as compared to the year 2019.

**Table 1:** Frequency and percentage change in Hospital admission and deaths before and during COVID 19 pandemic.

| **Admission parameters** | **2019**<br>**(Jan-June)** | **2020**<br>**(Jan-June)** | **Percentage**<br>**change** | **p-value** |
| --- | --- | --- | --- | --- |
| Total attendance (per month) | 23471 | 10903 | -54% | 0.02 |
| Routine OPD | 18971 | 8331 | -56% | .01 |
| PED ER attendance | 4500 | 2572 | -43% | 0.1 |
| Emergency admission | 1218 | 857 | -30% | 0.08 |
| Emergency Deaths<br>(Death within 24hrs of admission) | 10 | 6 | -35% | 0.17 |
| Death % | 0.8 | 1.2 | +56% | 0.38 |

**Figure 1:**
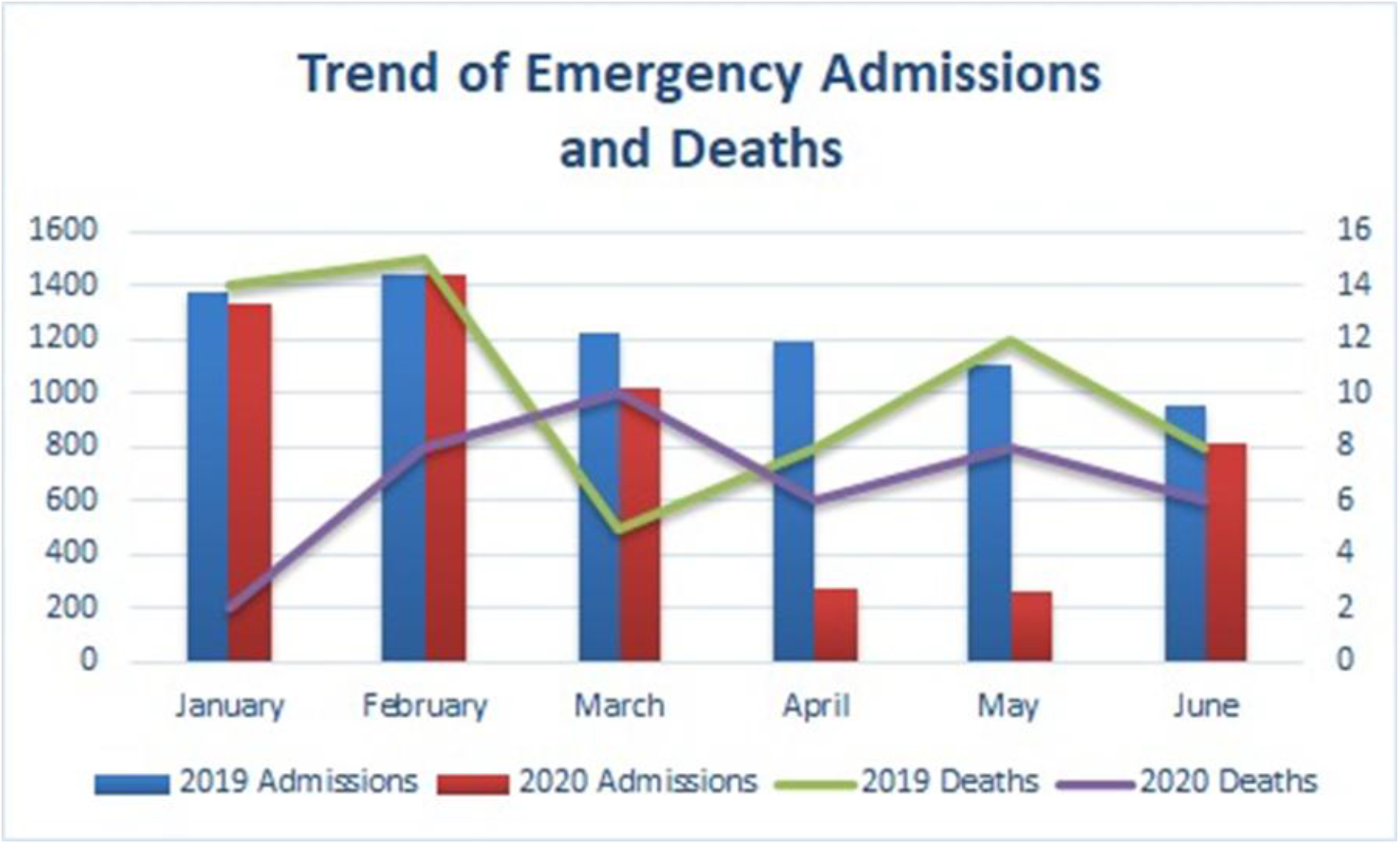
Trend of Emergency admissions and deaths.

### Impact of COVID 19 on disease categories in PED

Reduction in PED visits were noticed uniformly across all disease categories. However, the differences were not statistically significant.(Table 2) However, during the period of strict lock down (April – June 2020) a significant difference was noted. The number of patients who were brought in cardio-pulmonary arrest showed a significant difference (p-0.015).

**Table 2:** Emergency Department Admissions categorized with percentage reduction in 2020 as compared with 2019.

| **Presenting Disorder** | **2019** | **2020** | **Percentage Change** | **p-value** |
| --- | --- | --- | --- | --- |
| Injury and Poisoning | 153 | 89 | -42% | 0.18 |
| Respiratory Illness | 5164 | 2880 | -44% | 0.29 |
| Gastrointestinal disorders | 515 | 437 | -15% | 0.64 |
| Nervous system disorders | 778 | 767 | -1% | 0.94 |
| Cardiovascular Disorders | 41 | 37 | -10% | 0.77 |
| Surgical conditions | 16 | 20 | 25% | 0.72 |
| Renal Disorders | 79 | 57 | -28% | 0.35 |
| Emergency/ Life threatening | 278 | 384 | 38% | 0.49 |
| Cardio –pulmonary arrest | 4 | 13 | 225% | 0.12 |
| Expiry within 24hrs | 33 | 28 | -15% | 0.7 |
| Medico legal Cases | 15 | 7 | -56% | 0.07 |

## Discussion

In the year 2020 total pediatric attendance reduced by 54%, and similarly PED attendance reduced by 43%. This fall in attendance was further exaggerated during the period of strict lockdown with 75% drop in April and 81% drop in May 2020. This trend was consistent across several countries as documented by Isba et al in UK with 33.8%^1^, Dann et al in Ireland with 50% ^2^, Ferraro et al in Argentina^3^ with 38.5%, and Lazzerini M et al in Italy with 75-80%^4^ decline in pediatric emergency attendances respectively. This fall in attendance in Argentina (38.5%) was reported to have been exaggerated further in the Month of April and May to nearly 88%.^3^ However interestingly in Pakistan even after 1 week of this continued lockdown, there was a slow but steady rise in outpatient clinic visits for the next 6 weeks.^5^

These results are largely attributable to the stringent lockdown imposed restrictions placed on movement, and fear of COVID-19 resulted in children being brought in overly critical conditions. Furthermore, strict segregation of patients with flu like symptoms from non-suspected COVID-19 (from April 2020) and reduction in non-serious emergency visits mostly due to parental anxiety are some additional factors that could possibly explain these reductions.

Reduction in PED visits were noticed uniformly in all disease categories We observed an interesting change in patient profile, neonatal PED visits increased by 144%, and a marked decrease in PED visits of children in age group 1-5years. This increase in neonatal PED visits could probably be because most newborns were home-delivered and therefore did not receive appropriate perinatal care. On the contrary, significant (P-0.005) reduction in children could be due to the fact that children with respiratory illnesses were seen in a separate Flu OPD.

Incidence of Medico legal cases such as Poisoning and Injuries reduced by 56% probably because of strict home isolation and increased parental supervision. Similar observation in reduction of such cases was made by Lisa et al in Ireland.^2^

On analyzing the data for mortality, we found that the patients who presented in PED were in an extremely sick condition by the time they were attended by a healthcare worker. Mortality rate in May 2020 had gone up by nearly 3 times as compared to average monthly mortality in 2019 (Pre-COVID). This was probably because patients were brought to emergency in later stages of disease progression and in cardio-pulmonary arrest. In the process of steering clear of preventable morbidities and deaths, understanding this phenomenon of delayed visits is key in designing preventive strategies.^5^

While children might not have been directly affected by the COVID-19 pandemic, the ripple effect has indirectly caused more damage than anticipated. The poor health system support network caused unprecedented damage to the health of children because of sequelae including reemergence of vaccine preventable diseases, flaring up of diseases like tuberculosis and poor health outcomes because of interruption in treatment of chronic illnesses like thalassemia, HIV and pediatric malignancies. All this may lead to increased morbidity and mortality in children in future – not because of COVID 19 itself, but rather as an after effect. Neglect at the patient’s end – fueled by their constrained physical mobility – can have adverse repercussions on the social, psychological, and economic wellbeing of affected families.

## Data Availability

None

## Contribution

Conceptualization: SG, Data collection and analysis: RT, writing, review and editing: SG and VK.

## Ethic approval

The approval was taken from Institutional ethical committee of Lady Hardinge Medical College, New Delhi.

## What is already known on this topic?

➢ There is a decline in pediatric emergency department visits since the start of the COVID - 19 pandemic
➢ Children are the major sufferers

## What this study adds?

➢ With the onset of COVID-19 pandemic, infants and children in all age group suffer especially the newborns.
➢ During this period there is an increased risk of children been brought critically sick or in cardio respiratory arrest state.
➢ The maximum impact on pediatric emergency services was seen during the strict Lock down period

